# Wearable sensor and machine learning accurately estimate tendon load and walking speed during immobilizing boot ambulation

**DOI:** 10.1101/2023.06.03.23290612

**Authors:** Michelle P. Kwon, Todd J. Hullfish, Casey J. Humbyrd, Lorraine A.T. Boakye, Josh R. Baxter

**Affiliations:** University of Pennsylvania

**Keywords:** Musculoskeletal Loading, Long-Term Monitoring, CAM Boot

## Abstract

Achilles tendon injuries are treated with progressive weight bearing to promote tendon healing and restore function. Patient rehabilitation progression are typically studied in controlled, lab settings and do not represent the long-term loading experienced during daily living. The purpose of this study is to develop a wearable paradigm to accurately monitor Achilles tendon loading and walking speed using low-cost sensors that reduce subject burden. Ten healthy adults walked in an immobilizing boot under various heel wedge conditions (30°, 5°, 0°) and walking speeds. Three-dimensional motion capture, ground reaction force, and 6-axis inertial measurement unit (IMU) signals were collected per trial. We used Least Absolute Shrinkage and Selection Operator (LASSO) regression to predict peak Achilles tendon load and walking speed. The effects of using only accelerometer data, different sampling frequency, and multiple sensors to train the model were also explored. Walking speed models outperformed (mean absolute percentage error (MAPE): 8.41 ± 4.08%) tendon load models (MAPE: 33.93 ± 23.9%). Models trained with subject-specific data performed significantly better than generalized models. For example, our personalized model that was trained with only subject-specific data predicted tendon load with a 11.5 ± 4.41% MAPE and walking speed with a 4.50 ± 0.91% MAPE. Removing gyroscope channels, decreasing sampling frequency, and using combinations of sensors had inconsequential effects on models performance (changes in MAPE < 6.09%). We developed a simple monitoring paradigm that uses LASSO regression and wearable sensors to accurately predict Achilles tendon loading and walking speed while ambulating in an immobilizing boot. This paradigm provides a clinically implementable strategy to longitudinally monitor patient loading and activity while recovering from Achilles tendon injuries.

## Introduction

Tendon loading during rehabilitation following musculoskeletal injuries and/or surgical treatment impacts healing outcomes. Recent advances in orthopaedic mechanobiology directly link tissue loading with tissue healing outcomes.^1–3^ However, our limited capacity to quantify tissue loading in the real world restricts our ability to improve rehabilitation strategies aimed at promoting tissue healing. Achilles injuries demonstrate the link between tissue loading, injury, and healing outcomes. The loading mechanisms of these injuries vary from repetitive sub-maximal loads which result in tendinopathies to single supra-maximal loads which result in ruptures.^4,5^ While overloading and underloading can detrimentally affect the tendon, mechanical loading is also used as a clinical intervention. Safely introducing loading after an Achilles tendon injury promotes healing and leads to improved outcomes compared to prolonged non-weightbearing and/or immobilization.^6^ The current rehabilitation standard for surgical Achilles tendon interventions such as an Achilles repair and debridement is progressive tendon loading in an immobilizing boot. We know these boots reduce Achilles tendon loading,^7,8^ but strategies to optimize loading that maximize functional outcomes remains unclear. Continuously monitoring tendon loading while patients wear an immobilizing boot is necessary to improve rehabilitation guidelines.

Monitoring tendon loading during early rehabilitation stages is currently done using a lab-grade instrumented insole. Our group developed a physics-based algorithm that accurately quantifies Achilles tendon loading while ambulating in immobilizing boots (8). However, this technique is impractical for monitoring large patient populations because it relies on expensive, complex sensors that require daily patient involvement to charge and log the data. Instead, we propose developing a new paradigm to estimate tendon loading in immobilizing boots that is less burdensome but maintains similar accuracy.

Inertial measurement units (IMUs) are ideal because they are easy to use, low maintenance, low-cost, and ubiquitous in personal smart devices. IMUs do not directly measure interaction loads needed to estimate tendon loading but have other advantages that make them worth exploring as a scalable option for monitoring patient recovery. These advantages include resistance to dust and water (some IMUs have excellent dust and water resistance with ratings of IP68), long battery life (some IMUs can record weeks of data on a single charge), and minimal handling needs (some IMUs log data with no additional buttons for subjects to press). These IMU attributes are in many commercially available sensors and align with efforts to minimize burden to both users and providers. We expect that the minimized burden of these sensors will increase data capture rates, an essential component to monitoring. While IMU measurements are difficult to interpret, machine learning approaches have been used to transform linear accelerations and angular velocities into physiologically meaningful metrics.^9–11^ Recent studies have shown good machine learning performances when IMUs are used to predict musculoskeletal outcomes and biomechanical measurements like joint kinematics,^12–14^ but none, to our knowledge, have used only IMUs to predict Achilles tendon loading.

The purpose of our study is to develop an IMU-based monitoring paradigm that accurately estimates Achilles tendon loading and walking speed when ambulating in an immobilizing boot. This is a necessary step towards effectively monitoring real-world Achilles tendon loading in patients recovering from injuries and surgeries. We measured lower leg linear accelerations and rotational velocities using an IMU secured to the lateral aspect of an immobilizing boot. We then used these measurements to train and validate *generalized* models to predict peak Achilles tendon loading and walking speed during gait. We trained *hybridized* and *personalized* models which include subject-specific data in the training set. We also tested the effects of different data streams, sampling frequencies, and IMU placements to determine strategies that minimize burden while maximizing model accuracy. We hypothesize that our tendon load and walking speed models will have acceptable predictions, which we defined as under 20% mean absolute percentage error (MAPE). We expect that these findings will guide researchers and clinicians on how to implement wearable sensors in clinical populations.

## Methods

### Data Collection

We recruited ten subjects (3 females, age: 25 ± 2.4 yrs, BMI: 23.9 ± 6.56) to walk in our lab in an immobilizing boot across a range of walking speeds, stride patterns, and immobilization angles. We recruited healthy young adults who could comfortably walk 10.5 meters spans for 1 hour. Subjects had no previous or current Achilles injury or pain that would affect their gait and provided written informed consent in this study which was approved by our university’s Institutional Review Board.

Subjects wore laboratory clothing and an immobilizing boot (Formfit® Walker Air CAM, Ossur, Reykjavík, Iceland), commonly worn by patients recovering from Achilles tendon injuries, on their right side (**Figure 1**). Each subject wore a commercially available IMU (Opal, APDM, Portland, OR) attached to the lateral shank of the immobilizing boot. They wore additional IMUs on their left wrist, left lateral shank, and left foot. We acquired accelerometer data with a measurement range of ± 16 g at 100 Hz and gyroscope data with a measurement range of ± 2000 deg/s at 100 Hz. We specifically used the Opal IMUs to synchronously collect IMU data with other laboratory equipment. We measured Achilles tendon loading and walking speed using validated techniques to generate target values necessary to train and validate our predictive models. We measured plantar loads at a 100 Hz using a 3-part instrumented insole (Loadsol, Novel, St. Paul, MN) placed in the immobilizing boot. We measured walking speed by tracking the pelvis using an 8-camera markerless motion capture system (OptiTrack, Corvallis, OR). We synchronously collected IMU, instrumented insole, and motion capture data while subjects walked at 4 different walking speeds and 3 ankle immobilization angles. Subjects were instructed to walk 8 meters in the lab at 4 self-selected speeds: pathological, slow, medium, and fast. Pathological gait was prompted to the participant as “If there was pain in your right foot, how might you walk?”. Slow was explained as “a slower than average walking speed”. Medium was described as their “average, everyday walking pace.” Fast was instructed as a “If you were in a hurry, but still walking”. We tested each of these walking speeds across 3 boot configurations that are clinically used to progressively increase tendon loading: a 2.5” heel wedge to approximate 30º, a 0.5” heel wedge to approximate 5º, and no heel wedge to approximate 0º or neutral ankle position. We acquired data during 8-16 walking trials for every test condition, which generated 6,300 total strides in our entire experiment.

**Figure 1.**
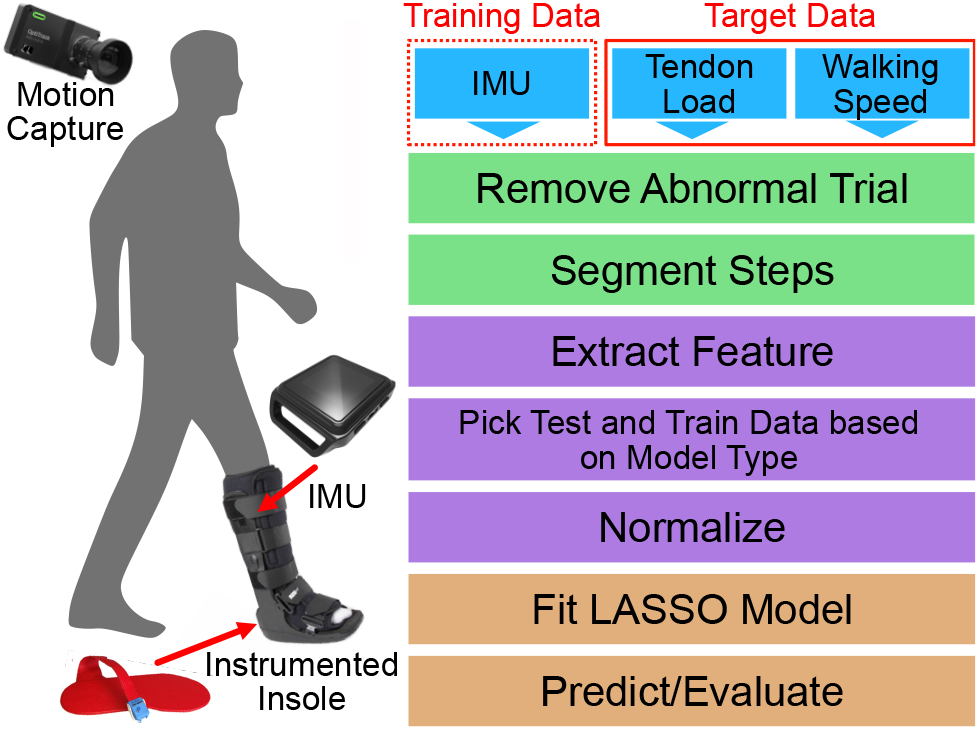
Flowchart of Methods. IMU, tendon load, and walking speed data were collected and serve as inputs (blue). The inputs are cleaned (green) and then pre-processed (purple). We then trained and tested our machine learning models (orange).

### Data Processing

We analyzed our experimental data using scientific computing software and custom scripts (python v3.9.12). We manually confirmed data integrity and rejected trials that were not time synchronized. We parsed data to isolate each gait cycle from heel strike to heel strike that we determined from the insole force data by setting a threshold of 50 N and then finding the local minima. For every gait cycle, the maximum, minimum, and impulse values of the IMU data in the corresponding period were calculated for all six channels (the superior-inferior, anterior-posterior, and lateral-medial directions for accelerometer and gyroscope). We estimated Achilles tendon loading using our validated instrumented insole paradigm^7,15^ and normalized these estimates to body weight (BW). Maximum tendon load was calculated and values under 0.5 BW were not included in the model given that surgical repair strength exceeds this threshold.^16^ Stance and swing times were also calculated through the force sensor. We tracked the body center of mass using markerless motion capture (Theia, Kingston, Ontario, Canada) and found the average walking speed by calculating the net forward progression of the center of mass divided by the time it took to traverse the space on a per trial basis. We confirmed that subjects walked at a constant speed throughout the trial with strong correlations (R^2^>.98) between center of mass displacement and time.

### Machine Learning Model

We selected Least Absolute Shrinkage and Selection Operator (LASSO) Regression to develop a predictive model for walking speed and peak Achilles tendon load. LASSO is a linear regression with an L1 regularization parameter and was implemented with the scikit-learn package (see Supplemental Material). We selected LASSO because it has been shown to successfully predict biomechanical proxies including joint kinematics and tibial loading.^12,17^ It has also outperformed other models including convoluted neural network and Gaussian Process Regressions.^11^ LASSO also is recommended to reduce feature dimensionality and minimize multicollinearity effects.^18,19^

We used maximum, minimum, absolute impulse, impulse of swing phase, and impulse of the stance phase of all IMU channels, along with stance and swing time for each heel strike to heel strike step as features and scaled them appropriately. Maximum Achilles tendon load and walking speed were used as prediction targets. We selected peak tendon loading as one of our models’ target measures because it is the most relevant biomechanical measure for Achilles tendon injuries and therefore, the desired metric for long-term monitoring. We also predicted walking speed because walking speed is a clinically advisable metric and is more likely to be better predicted by IMU data. We also expect walking speed and Achilles tendon load to be positively correlated where faster walking speeds lead to higher tendon load. Thus, recommending patients to adjust their walking speed may allow clinicians to prescribe a specific tendon load during rehabilitation.

We developed separate models for each immobilizing boot condition (30º, 5º and 0º) because progressively removing heel wedges is a key element in the protocol that clinicians prescribe and results in fundamentally different tendon loads. Each model was trained through a k-fold cross validation method where the IMU data was split into 10 partitions, 1 for every subject. One partition (a test subject) was excluded to test the model while the remaining 9 were used to train a model. Because the training data of this model did not incorporate any of the test subject’s data, we considered this model to be made with a *generalized strategy*. We used the MAPE as the evaluation metric and set an *a priori* ‘excellent’ agreement threshold of 10% MAPE and ‘acceptable’ agreement threshold of 20% MAPE. Each subject partition was excluded once and the average of the MAPEs were reported to assess the robustness of the model.

We then explored incorporating subject-specific data into the model to improve performance. Unlike many other machine learning settings, we can produce new subject data during lab or clinic visits using our instrumented insole paradigm or simple walking speed measurements. These data could be incorporated into the model to make tailored predictions unique for that individual. We tested two different methodologies aimed at improving model performance. In the *hybridized strategy*, we incorporated 50% of one subject’s data to the other 9 subjects’ data to form the training dataset and tested the model on the remaining 50% of data. In the *personalized strategy*, we solely trained a model on 50% of one subject’s data and tested the model’s performance on the remaining 50% of that one subject’s data. These 50% of a subject’s data equated to roughly 82 steps, which is a small fraction of the steps taken during daily living and is feasible to collect during lab or clinical visits.

We performed additional analyses to test how excluding the gyroscope data stream, different sampling frequencies, and multiple sensors impact model performance. These are important considerations because we want to decrease user burden by maximizing battery life while minimizing the number of wearable sensors. By recording only accelerometer, the battery life of some IMU-based wearables can double. Thus, we also trained models without the gyroscope to assess their performance of predicting tendon load and walking speed against models trained with both accelerometer and gyroscope data. Decreasing the sensor’s sampling frequency further extends IMU battery, so we assessed the performance of models trained on data that was resampled to simulate 50 Hz and 25 Hz data. Eliminating the gyroscope data and decreasing the sampling frequency are expected to impact model performance negatively, so we tested the impact of training models with multiple sensors, which we expect would increase model performance. We did this by assessing the effects of including data from IMUs on the contralateral limb, including the contralateral wrist, contralateral shank, and contralateral foot (**Figure 1**).

## Results

Walking speed predictions were consistently better than the Achilles tendon load predictions. When predicting tendon load using the IMU data, the MAPE values were highest in the 5º condition at 44.0% and lowest in 0º at 25.8%. In the walking speed models, the highest MAPE was in the 30º condition at 10.6% and lowest in the 0º at 6.20% (**Tables 1 and 2**). The most important features in the Achilles tendon load model were the gyroscope in the lateral-medial direction, the time of stance and swing, and the stance phase impulse of the accelerometer in the superior-inferior direction. For the walking speed models, similar features played an important role including the impulse of the accelerometer in the superior-inferior direction and the stance and swing time.

**Table 1.** MAPE values of Tendon Load and Walking Speed by Model Paradigm Type (2 columns)

| Boot Condition (deg) | Tendon Load MAPE (%) |  |  | Walking Speed MAPE (%) |  |  |
| --- | --- | --- | --- | --- | --- | --- |
|  | Generalized | Hybridized | Personalized | Generalized | Hybridized | Personalized |
| 30 | 31.9 ±19.9 | 22.7 ±9.34 | 10.7 ±2.94 | 10.6 ±4.83 | 8.26 ±3.14 | 5.35 ±0.999 |
| 5 | 44.0 ±31.0 | 26.4 ±19.7 | 12.6 ±5.07 | 8.42 ±4.48 | 5.16 ±1.60 | 3.69 ±0.882 |
| 0 | 25.8 ±20.8 | 19.6 ±13.0 | 8.35 ±2.90 | 6.20 ±2.92 | 4.51 ±1.38 | 3.38±0.816 |

Reported as MAPE ±STD

**Table 2.** MAPE values of Tendon Load and Walking Speed predicted using Generalized Models with Adjusted Parameters (2 columns)

| IMU Channels | Sampling Frequency (Hz) | Location | Boot Condition (deg) | Tendon Load MAPE (%) | Walking Speed MAPE (%) |
| --- | --- | --- | --- | --- | --- |
| accelerometer + gyroscope | 100 | Boot | 30 | 31.9 ±19.9 | 10.6 ±4.83 |
|  |  |  | 5 | 44.0 ±31.0 | 8.42 ±4.48 |
|  |  |  | 0 | 25.8 ±20.8 | 6.20 ±2.92 |
| accelerometer | 100 | Boot | 30 | 49.3±24.5 | 9.94±3.41 |
|  |  |  | 5 | 42.0±36.6 | 8.09± 3.28 |
|  |  |  | 0 | 28.8±22.6 | 9.20±4.09 |
| accelerometer + gyroscope | 50 | Boot | 30 | 34.6±21.8 | 11.5±5.27 |
|  |  |  | 5 | 34.3±22.9 | 9.62±4.65 |
|  |  |  | 0 | 24.4±21.3 | 7.15±3.27 |
| accelerometer + gyroscope | 25 | Boot | 30 | 36.5±21.4 | 12.0±5.49 |
|  |  |  | 5 | 35.9±25.6 | 10.1±5.05 |
|  |  |  | 0 | 28.2±24.8 | 9.84±4.91 |
| accelerometer + gyroscope | 100 | Boot + Left Foot | 30 | 36.6±25.1 | 8.66±3.95 |
|  |  |  | 5 | 42.1±35.5 | 5.13±1.68 |
|  |  |  | 0 | 32.4±20.9 | 5.37±2.39 |

Reported as MAPE ±STD

We found using subject-specific data to train the model resulted in better predictions with decreased MAPE scores. When incorporating 50% of the subject’s data to train hybridized models, we found that MAPEs decreased on average by 11.0% for the tendon load models and 2.43% for walking speed models. When training personalized models using subject-specific data, we observed larger improvements in MAPEs. The average decrease in MAPEs when compared to the generalized models are 23.4% for Achilles tendon load and 4.27% for walking speed. The MAPEs of the personalized models were the lowest for both (**Table 1**) tendon load (**Figure 2**) and walking speed predictions (**Figure 3**).

**Figure 2.**
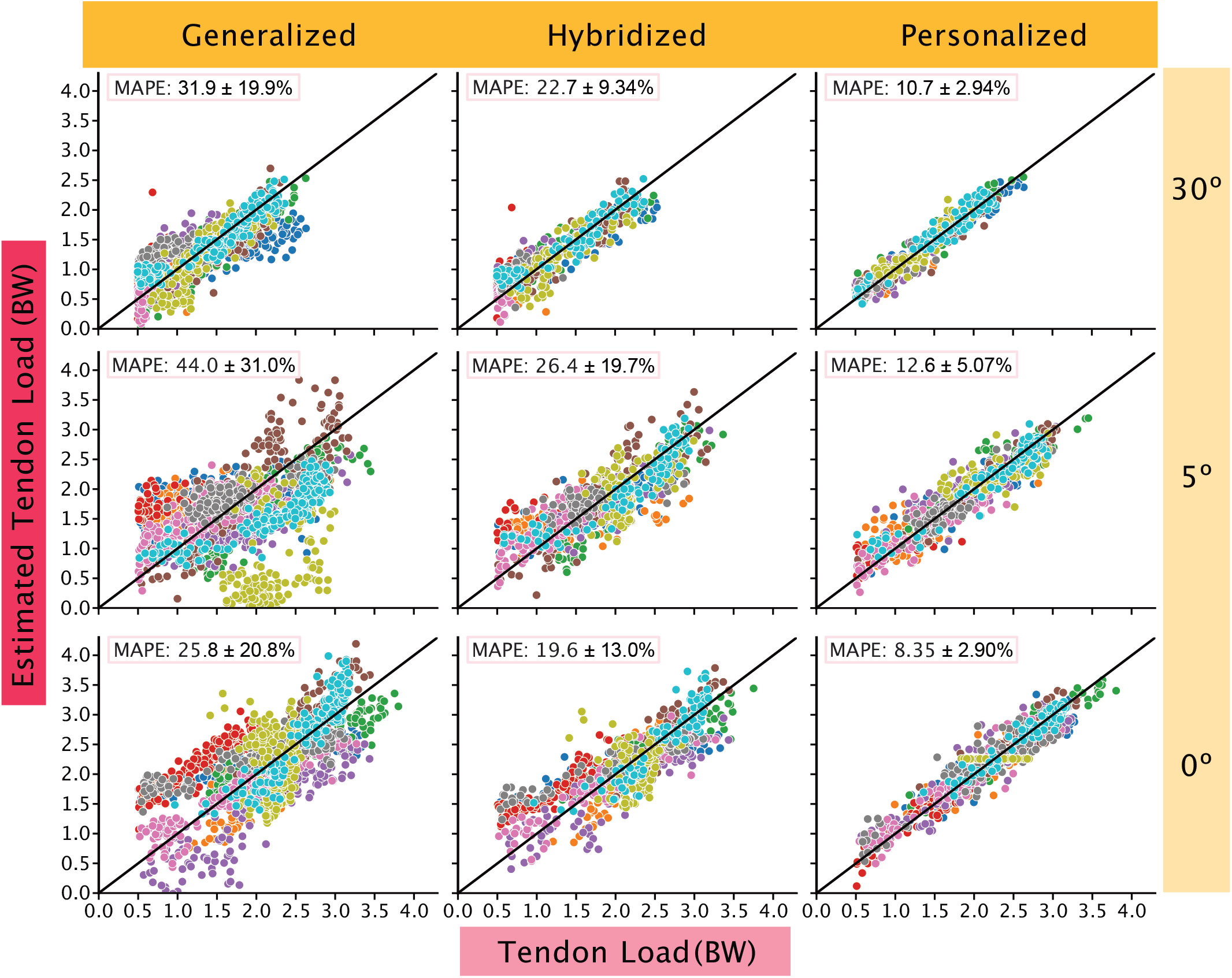
Tendon loading predictions improved when subject-specific data were added to the models. The generalized models (left column) were least accurate and tended to spread subject data (each subject data is represented by different colored circles) away from the reference line (black line), which represents 0 MAPE. We built hybridized models (center column) by adding subject-specific data to the generalized models that improved performance. However, personalized models (right column) were the most accurate at predicting tendon loading across all boot conditions and all fell in the ‘acceptable’ to ‘excellent’ category. Out of the three boot conditions, the 5º condition had the widest spread.

**Figure 3.**
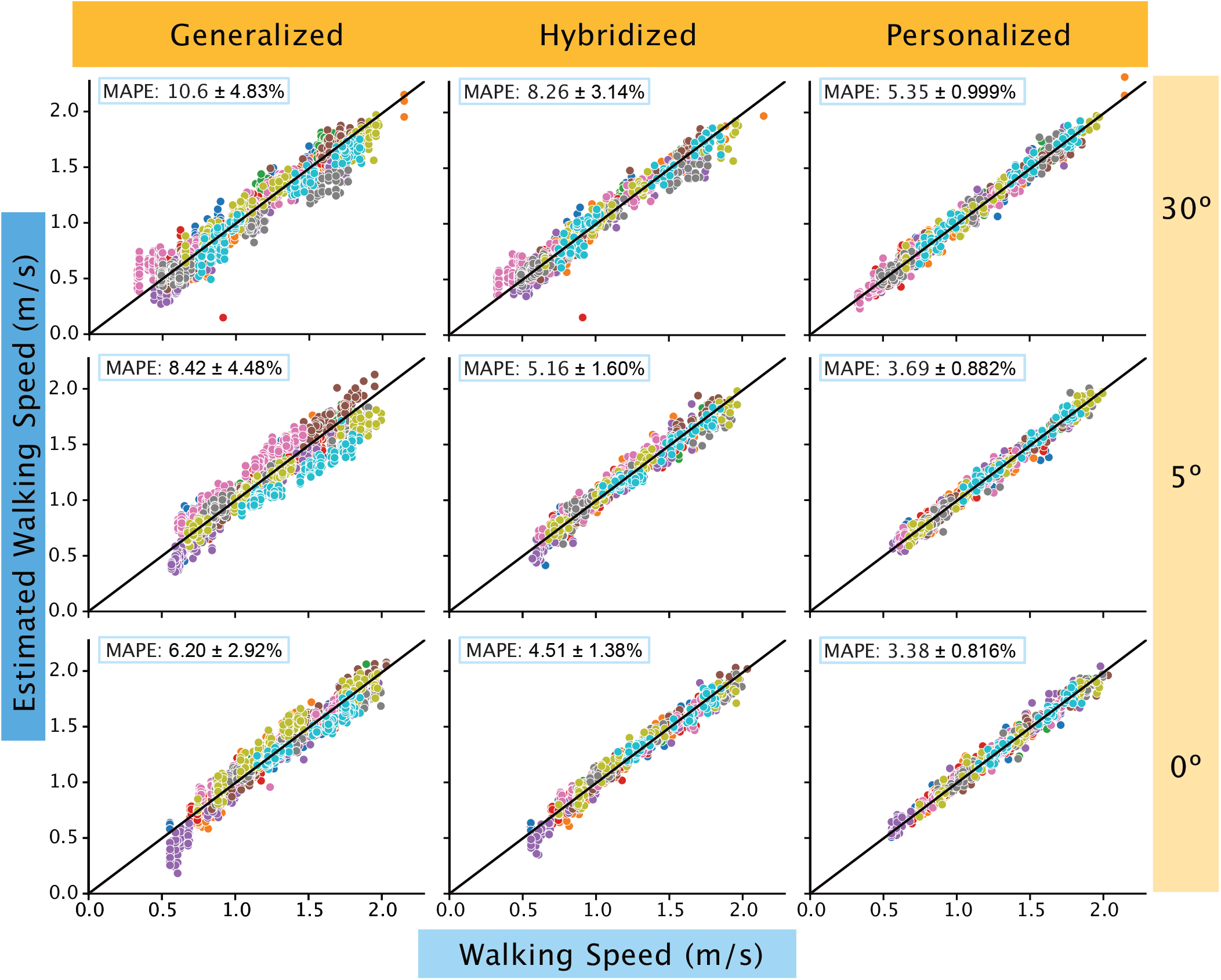
All walking speed predictions were within the ‘acceptable’ to ‘excellent’ threshold. When subject specific data was added to the generalized model (left column), the hybridized model (center column) were ‘excellent’ in prediction. The personalized models (right column) further improved in accuracy as evidenced by the close clustering of all subject data (each subject data is represented by different colored circles) towards the reference line (black line). The walking speed predictions consistently improved with progressive loading with 30º (top row) having the widest spread away from the reference line compared to 0º (bottom row).

Training the tendon load and walking speed models without gyroscope data had varying effects on model performance. When predicting Achilles tendon load, excluding gyroscope data had minimal effects on model performance decreasing the 5º condition MAPEs by 2.06% and increasing 00º condition MAPEs by 2.99%. However, the 30º tendon load model experienced a drastic increase in MAPE by 17.34%. We observed small changes in the walking speed models when excluding gyroscope data which resulted in <1% decreases for the 30º and 5º boot condition and a 3.01% increase in the 0º boot condition. Despite the varying effects on model performance, excluding gyroscope data did not ultimately change the overall usability of these models. All tendon load models continued to not meet ‘acceptable’ threshold (MAPE <20%) and all walking speed models met the ‘excellent’ MAPE threshold (MAPEs <10%) (**Table 2**).

New models for tendon load and walking speed trained with 50 Hz and 25 Hz data did not materially impact model performance. For the tendon load models, all MAPEs remained above 20%. However, based on the sampling frequency and condition, tendon load MAPEs either increased or decreased when compared to models trained on 100 Hz data. For example, the 0º condition, tendon load model’s MAPE decreased to 24.7% when using 50 Hz data but increased to 27.7% when using 25 Hz data. However, both changes are marginal. While the changes between 100 Hz, 50 Hz, and 25 Hz did not consistently increase or decrease, the models trained with the 25 Hz data were consistently worse with increased MAPEs when compared to the 50 Hz data. Unlike the tendon load models, walking speed MAPEs for all conditions for 50 Hz data increased the average MAPE by .541% and 25 Hz increased the average MAPE by 1.87% when compared to the MAPEs of models trained on 100 Hz data. Similar to the tendon load models, the walking speeds produced higher MAPEs when trained on 50 Hz data than when predicting with 25 Hz data (**Table 2**).

Lastly, combining data from a sensor on the boot with data from sensors on the contralateral extremities had minimal effects on model performance. For the models that predict tendon load, no combination of sensor helped improve the predictive power across the 30º, 5º, and 0º immobilizing boot conditions. When looking at average of sensor combination effects across the three different boot conditions, all MAPEs increased when compared to only using the sensor on the lateral boot. For walking speed, the most favorable sensor combination was training the model with data from the boot and the contralateral foot, which had MAPEs of 8.66% for the 30º, 5.13% for 5º, and 5.37% for 0º condition (**Table 2**). All other MAPE values for their respective boot sensor combination are reported as a supplement (**see Supplementary Material**).

## Discussion

We developed a robust LASSO regression model that uses data from a commercially available IMU secured to an immobilizing boot to predict peak Achilles tendon loads and walking speeds with high levels of confidence. The predictions of the generalized and hybridized models did not fall within the ‘acceptable’ range for Achilles tendon loading. However, the personalized tendon load models were all considered ‘acceptable’ with the 0º tendon load model even being ‘excellent’. For walking speed predictions using the generalized models, the 5º and 0º hit our ‘excellent’ threshold. However, the 30º condition model fell shortly out of the <10% range with a 10.6% MAPE. The hybridized and personalized walking speed models performed even better, all having ‘excellent’ predictive power.

Removing the gyroscope data stream, reducing sampling frequency, and the altering IMU placement all had varying effects on the model. None altered the implementation potential for long-term monitoring contexts, ultimately making their effects unremarkable. Therefore, the models’ performance can be tuned by adjusting parameters including incorporating multiple sensors and battery life optimization (remove gyroscope and reduce sampling frequency) without altering MAPE values in a meaningful way. We recommend that researchers and clinicians adjust these parameters by carefully considering the tradeoffs between model performance and user burden.

Since the personalized Achilles tendon load models and generalized walking speed models both predict their target metrics with ‘acceptable’ to ‘excellent’ performance, an important clinical decision of which monitoring paradigm to deploy arises. One monitoring paradigm accurately predicts walking speed without collecting subject specific data using the generalized model, which eliminates the need for an in-lab visit before monitoring. The other monitoring paradigm to consider is accurately predicting Achilles tendon load using a personalized model, which would require an in-lab visit to collect approximately 82 steps. However, we do not expect these steps to be difficult to obtain. Some possible ways to collect the data is a 5-minute lab visit, walking around the clinic with the sensors for 5 minutes, or requesting that the patient self-monitors with the sensors for one day.

The first consideration to choose the appropriate monitoring paradigm is to consider whether it is possible and worthwhile to obtain subject-specific training data. To collect training data, one must have the time (approx. 5 minutes), space (lab space or a long hallway to collect continuous walking data), and materials (instrumented insole) to conduct a brief data collection session. These data will later be used to create the personalized model to predict tendon load. The next consideration to choose the appropriate monitoring paradigm to implement is whether the preferred biomechanical metric is walking speed or tendon load. Walking speed is clinically useful because it is a metric that is easy to understand and modify through coaching. For example, people can more easily alter their walking speed than the amount of load going through their Achilles tendon. However, Achilles tendon load is a more direct measurement, which may be a better indicator when trying to optimize functional outcomes. Thus, researchers and clinicians should consider what metric would best accomplish their goals (**Figure 4**). Another factor to consider is what MAPE value can be tolerated. Our generalized walking speed model (**Figure 3**) is better suited for situations where error values must be minimized because it performed better than the tendon loading models – even models developed using personalized data (**Figure 2**).

**Figure 4.**
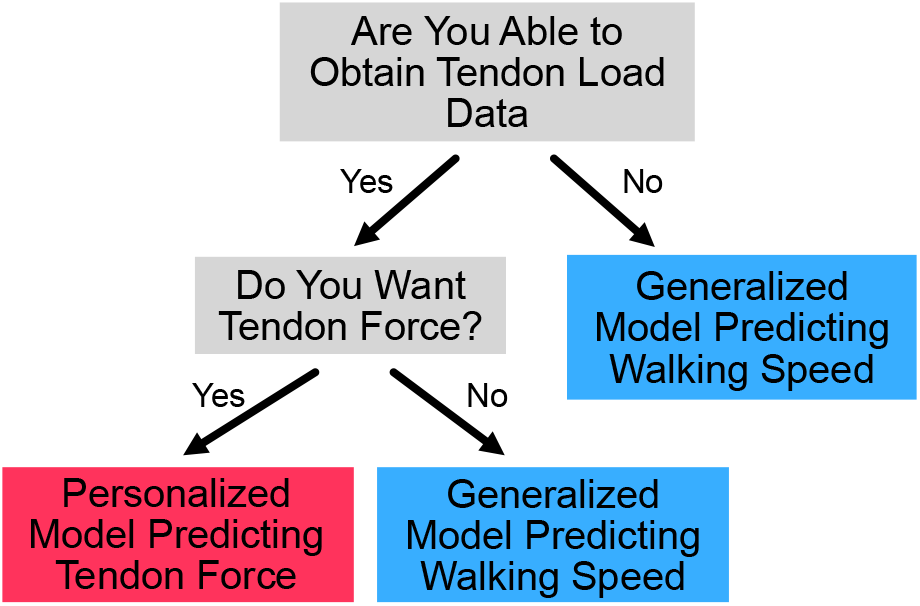
Decision tree to select appropriate monitoring paradigm to deploy based on clinician/researcher needs. Users should consider whether tendon loading predictions are necessary for the research question or clinical feedback and whether tendon loading data can be obtained to train patient specific models.

In addition to selecting an appropriate monitoring paradigm, minimizing user burden is also an important factor to consider when designing longitudinal studies that monitor subject activity or loading patterns. One way to decrease burden is to increase sensor battery life, which can be accomplished by using only accelerometry functionality on the IMU or collecting data at a lower sampling frequency. For example, collecting only accelerometer data and decreasing the sampling frequency to 50 Hz can quadruple the battery life, allowing some IMUS to record for 34 days on a single charge.^20^ Maximizing battery life allows us to continuously collect patient data between clinical checkups following Achilles tendon rupture with minimal patient burden. Our results confirm that training the model using 50 Hz data and no gyroscope data to maximize battery life decreases the MAPE of the generalized tendon load model by 5.87% on average but increases the MAPE of the generalized walking speed model by 1.00% on average. For personalized models the effect of decreasing sampling frequency and removing gyroscope is smaller. Training on data without gyroscope and at 50 Hz increased the average MAPE of personalized tendon load predictions by 1.33% and of personalized walking speed predictions by .783%. Given the minimal increases in MAPE in both the generalized models and the personalized models when removing the gyroscope and sampling at 50 Hz, we consider this increased MAPE a worthwhile tradeoff to quadruple the battery life.

We identified two promising monitoring paradigms that balance the tradeoffs between model performance and user burden (**Figure 5**). The first optimal paradigm is to train a personalized model that predicts tendon load using a single sensor on the immobilizing boot that samples at 50 Hz without gyroscope. Under this configuration, the 30º (MAPE = 12.0%) and the 5º condition (MAPE = 14.6%) were ‘acceptable’ (MAPE<20%). The model to predict tendon force in the 0º condition had ‘excellent’ predictive strength (<10%) with a MAPE of 9.22%. The second optimal paradigm is to train a generalized model that predicts walking speed using a single sensor on the immobilizing boot that samples at 50 Hz without gyroscope. The 0º (MAPE = 9.63%) and the 5º (MAPE = 8.18%) predictions were both ‘excellent’ (MAPE <10%) while the 30º (MAPE = 10.4%) prediction was ‘acceptable’. Therefore, both monitoring paradigms guarantee adequate predictions of a clinically relevant biomechanical metric while minimizing the burden placed on the user, making them ideal to implement for long-term monitoring contexts.

**Figure 5.**
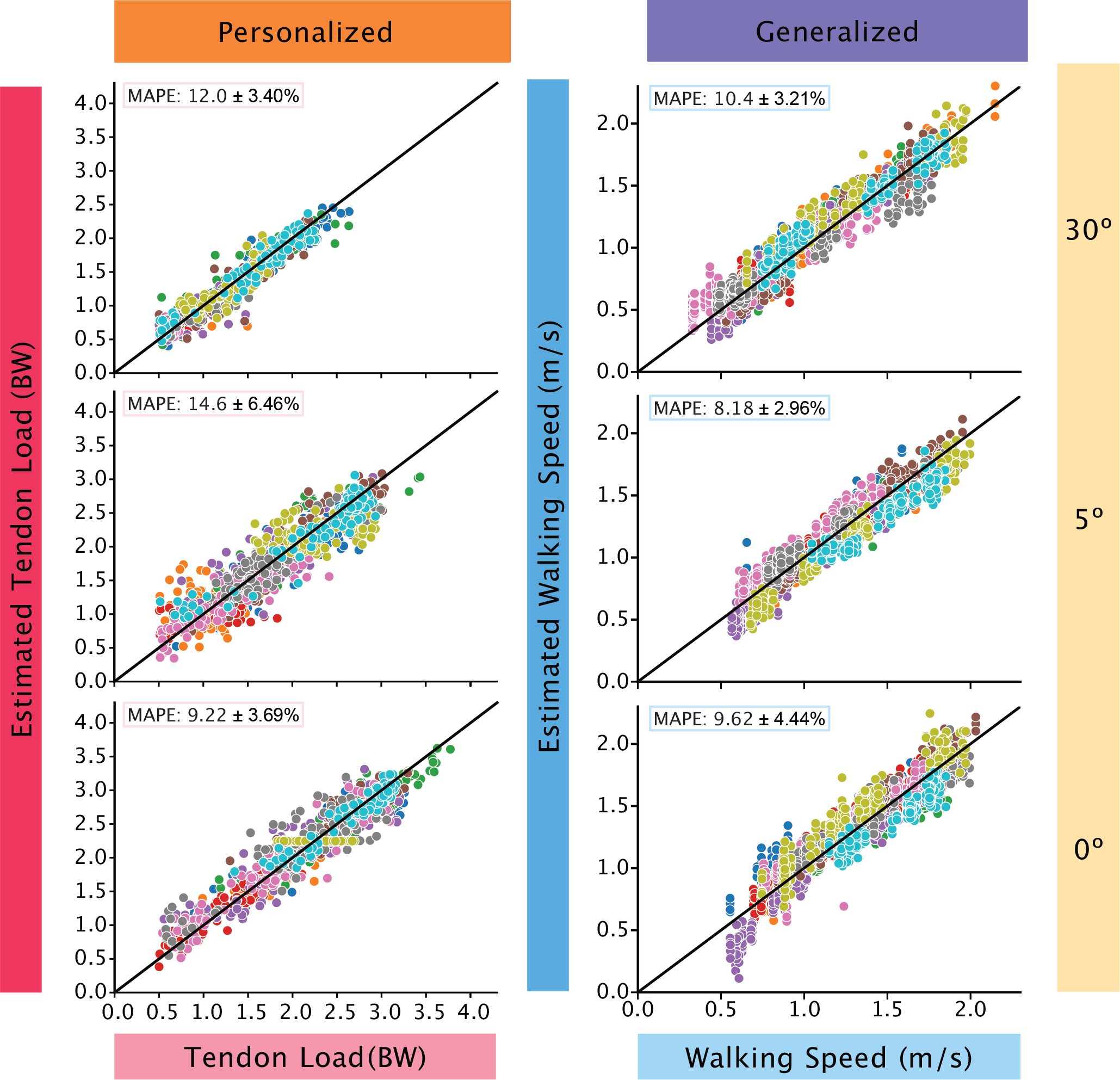
The model performance of two promising monitoring paradigms were all ‘acceptable’ or ‘excellent’. The left column depicts the model performance of estimating tendon load using a personalized model with data at 50 Hz with only accelerometer data. The right column shows the model performance of estimating walking speed using a generalized model with data at 50 Hz with only accelerometer data. The accurate predictions of both these monitoring paradigms allow us to monitor Achilles tendon health for long periods of time with minimal subject burden.

Commercially available IMUs are quickly becoming ubiquitous, but it is often difficult to identify an optimal sensor to deploy in the field. We identified 4 IMU criteria that we believe will maximize subject retention: 1) the battery life must exceed 1 week, 2) the sensor does not require daily interaction, 3) the sensor is resistant to dust and water experienced in the real-world, and 4) the sensor does not require additional components to use like a smartphone. We found a commercially available IMU (ax6, Axivity) that met our criteria. The accelerometer-only version of this IMU was used to continuously capture 1-week physical activity from 100,000 participants in the UK Biobank (22).^21^ During this study, we used APDM Opal IMUs to time synchronize IMU data with our laboratory equipment to measure Achilles tendon loading with our instrumented insoles and walking speed with motion capture. However, Axivity’s AX6 IMUs are better to deploy in-field. To ensure that Opal IMUs and Axivity IMUs would perform similarly, we performed a validation. In this validation experiment we secured both sensors to a rigid bar and tested the effects of gravity on the 3 accelerometer axes and swung the bar to approximate a range of boot motions. As expected, we found that the accelerations caused by gravity and the angular velocities while swinging between the 2 sensor types were strongly correlated with an average cross correlation score of 98.7% (**Figure 6**). This demonstrates that laboratory grade IMUs, like the Opal, can be used to develop training data sets for field ready IMUs, like the AX6, that are better suited for measuring activity patterns in the real-world. We want to stress that at the time of this study, we found that this specific IMU met our criteria. But we anticipate that other IMU solutions will quickly emerge as the wearable space in healthcare continues to improve.

**Figure 6.**
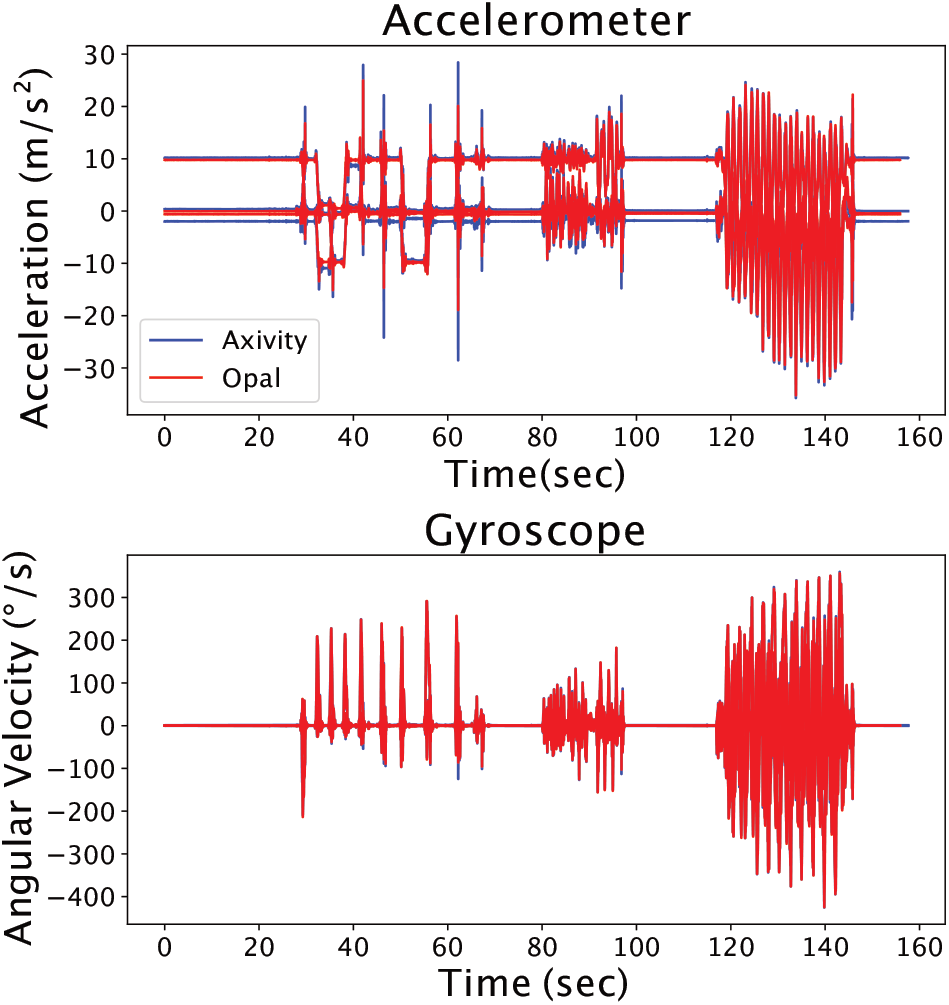
Axivity (blue) and Opal (red) IMU data show strong agreement. While these findings are expected, they confirm that IMU sensors can be selected based on selection criteria that satisfy the research or clinical need rather than IMU measurement fidelity.

There are various adjustments and improvements that can be made to optimize our models’ performance when predicting both tendon load and walking speed. Collecting data from more subjects will increase the models’ robustness by capturing more gait variations, but these gains are quickly diminishing. We calculated a learning curve from our data and found the validation curve to level off with more data. The average difference in MAPE between the validation curve when a model is trained on 80% and 90% of the dataset was 1.73%. The stability of the learning curve as more data was added and the small difference of MAPEs observed with larger training sizes suggests that collecting more data would not significantly alter MAPEs if more data were to be collected. We expect that walking speed was more consistently predictable between different people since it is governed by segment kinematics, and the model parameters, like stride pace and peak accelerations, were directly captured in accelerometry data. Therefore, once generalized walking speed models are well-trained, we expect these models to accurately predict new subject data. Conversely, muscle-tendon loads that drive motion are more variable between individuals and this will be compounded by physical constraints – like wearing an immobilizing boot – or injury and pain – like patients recovering from tendon injury.

The changes induced by the different physical constraints will have various manifestations in accelerometer and gyroscope data and do not affect each person in the same manner. Therefore, specific-subject data are needed to train personalized models that accurately reflect each subject’s unique tendon loading. Future work should focus on including other biomechanical signals to improve tendon load predictions like plantar loading or plantar flexor muscle activity. Recent work by Matijevich et al. demonstrated that machine learning algorithms predict musculoskeletal loading better when loading measures are included in the model (18). This is an important consideration before moving away from personalized models to predict tendon loading. However, the same tradeoff calculation we made between model performance and user burden need to be made when introducing new sensor types that come with their own limitations.

Our monitoring paradigm has several limitations that should be considered before implementing wearables in clinical research. We studied healthy controls with no relevant history of an Achilles injury. The LASSO models trained from their data do not capture the gait cycle abnormalities typically seen in subjects who are injured or recovering from surgery. We instructed our subjects to walk with a ‘pathologic’ gait, but we could not confirm that this gait was equivalent to patient gait. While the results from this study were a good estimate of the accuracies and errors that we may expect if we run the same models on patient training data, we admit that these models do not necessarily transfer to patients. Our study demonstrates proof-of-concept, but large patient data sets are needed to train LASSO models specific to Achilles injury patient populations.

We successfully developed two monitoring paradigms that make long-term monitoring of subjects feasible. One monitoring paradigm consists of creating unique, personalized models to predict tendon load, while the other paradigm consists of creating a generalized model to predict walking speed. Both models lead to highly accurate predictions of tendon load and walking speed. Each model should be further tuned based on the researcher’s or clinician’s needs by using different combinations of IMUs or choosing different settings. These monitoring paradigms allow for a cost-effective, long-term monitoring solution with minimal burden to the subject. This new capability to longitudinally monitor patients is a necessary tool that will allow us and other rehabilitation researchers to develop evidence-based rehabilitative loading strategies that optimize patient healing outcomes. By doing so, we will develop new evidence-based clinical recommendations that promote Achilles tendon healing.

## Data Availability

Complete results, analysis code, and study data are available online. Raw study data are available upon reasonable request.

https://upenn.box.com/v/boot-IMU-LASSO

## Acknowledgments

This work was funded by NIH/NIAMS P50-AR080581 and R01-AR078898.

## Supplemental Digital Content

All supplemental data are available at https://upenn.box.com/v/boot-IMU-LASSO

- Final_Result_Datasheet.xlsx – complete data results sheet
- final_code.py – python code that runs LASSO to train and evaluate models on experimental data
- dataset.pkl – python pickle file that contains all experimental data

## References

1. Jielile, J. et al. Early Ankle Mobilization Promotes Healing in a Rabbit Model of Achilles Tendon Rupture. Orthopedics 39, e117–126 (2016).

2. Malliaras, P., Barton, C. J., Reeves, N. D. & Langberg, H. Achilles and Patellar Tendinopathy Loading Programmes. Sports Med 43, 267–286 (2013).

3. Alfredson, H., Pietilä, T., Jonsson, P. & Lorentzon, R. Heavy-load eccentric calf muscle training for the treatment of chronic Achilles tendinosis. Am J Sports Med 26, 360–366 (1998).

4. Rompe, J. D., Furia, J. P. & Maffulli, N. Mid-portion Achilles tendinopathy--current options for treatment. Disabil. Rehabil. 30, 1666–1676 (2008).

5. Millar, N. L. et al. Tendinopathy. Nature Reviews Disease Primers 7, 1–21 (2021).

6. Merza, E., Pearson, S., Lichtwark, G., Ollason, M. & Malliaras, P. Immediate and long-term effects of mechanical loading on Achilles tendon volume: A systematic review and meta-analysis. J Biomech 118, 110289 (2021).

7. Hullfish, T. J., O’Connor, K. M. & Baxter, J. R. Instrumented immobilizing boot paradigm quantifies reduced Achilles tendon loading during gait. Journal of Biomechanics 109, 109925 (2020).

8. Smyth, N. A. et al. The Effect of CAM Boots on Contact Pressures of the Ankle and Hindfoot Joints. Foot Ankle Orthop 5, 2473011420S00014 (2020).

9. Lee, Y.-S. et al. Assessment of walking, running, and jumping movement features by using the inertial measurement unit. Gait Posture 41, 877–881 (2015).

10. Kwon, H., Abowd, G. D. & Plötz, T. Complex Deep Neural Networks from Large Scale Virtual IMU Data for Effective Human Activity Recognition Using Wearables. Sensors (Basel) 21, 8337 (2021).

11. Kammoun, N., Apte, S., Karami, H. & Aminian, K. Estimation of Temporal Parameters During Running with a Wrist-worn Inertial Sensor: an In-field Validation. Annu Int Conf IEEE Eng Med Biol Soc 2022, 3669–3672 (2022).

12. De Brabandere, A. et al. A Machine Learning Approach to Estimate Hip and Knee Joint Loading Using a Mobile Phone-Embedded IMU. Front Bioeng Biotechnol 8, 320 (2020).

13. Sharifi Renani, M., Eustace, A. M., Myers, C. A. & Clary, C. W. The Use of Synthetic IMU Signals in the Training of Deep Learning Models Significantly Improves the Accuracy of Joint Kinematic Predictions. Sensors (Basel) 21, 5876 (2021).

14. Chaaban, C. R. et al. Combining Inertial Sensors and Machine Learning to Predict vGRF and Knee Biomechanics during a Double Limb Jump Landing Task. Sensors (Basel) 21, 4383 (2021).

15. Hullfish, T. J. & Baxter, J. R. A simple instrumented insole algorithm to estimate plantar flexion moments. Gait & Posture 79, 92–95 (2020).

16. Demetracopoulos, C. a., Gilbert, S. L., Young, E., Baxter, J. R. & Deland, J. T. Limited-Open Achilles Tendon Repair Using Locking Sutures Versus Nonlocking Sutures: An In Vitro Model. Foot & Ankle International 35, 612–618 (2014).

17. Matijevich, E. S., Scott, L. R., Volgyesi, P., Derry, K. H. & Zelik, K. E. Combining wearable sensor signals, machine learning and biomechanics to estimate tibial bone force and damage during running. Human Movement Science 74, 102690 (2020).

18. Halilaj, E. et al. Machine learning in human movement biomechanics: Best practices, common pitfalls, and new opportunities. J Biomech 81, 1–11 (2018).

19. Kipp, K. & Warmenhoven, J. Applications of regularized regression models in sports biomechanics research. Sports Biomech 1–19 (2022) doi:10.1080/14763141.2022.2151932.

20. Axivity Ltd. AX6 Datasheet. 6-Axis logging movement senso. (2023).

21. Data-Field 90001. https://biobank.ctsu.ox.ac.uk/crystal/field.cgi?id=90001.

